# Integrative analysis of clinical health records, imaging and pathogen genomics identifies personalized predictors of disease prognosis in tuberculosis

**DOI:** 10.1101/2022.07.20.22277862

**Authors:** Awanti Sambarey, Kirk Smith, Carolina Chung, Harkirat Singh Arora, Zhenhua Yang, Prachi Agarwal, Sriram Chandrasekaran

## Abstract

Tuberculosis (TB) afflicts over 10 million people every year and its global burden is projected to increase dramatically due to multidrug-resistant TB (MDR-TB). The Covid-19 pandemic has resulted in reduced access to TB diagnosis and treatment, reversing decades of progress in disease management globally. It is thus crucial to analyze real-world multi-domain information from patient health records to determine personalized predictors of TB treatment outcome and drug resistance. We conduct a retrospective analysis on electronic health records of 5060 TB patients spanning 10 countries with high burden of MDR-TB including Ukraine, Moldova, Belarus and India available on the NIAID-TB portals database. We analyze over 200 features across multiple host and pathogen modalities representing patient social demographics, disease presentations as seen in cChest X rays and CT scans, and genomic records with drug susceptibility features of the pathogen strain from each patient. Our machine learning model, built with diverse data modalities outperforms models built using each modality alone in predicting treatment outcomes, with an accuracy of 81% and AUC of 0.768. We determine robust predictors across countries that are associated with unsuccessful treatmentclinical outcomes, and validate our predictions on new patient data from TB Portals. Our analysis of drug regimens and drug interactions suggests that synergistic drug combinations and those containing the drugs Bedaquiline, Levofloxacin, Clofazimine and Amoxicillin see more success in treating MDR and XDR TB. Features identified via chest imaging such as percentage of abnormal volume, size of lung cavitation and bronchial obstruction are associated significantly with pathogen genomic attributes of drug resistance. Increased disease severity was also observed in patients with lower BMI and with comorbidities. Our integrated multi-modal analysis thus revealed significant associations between radiological, microbiological, therapeutic, and demographic data modalities, providing a deeper understanding of personalized responses to aid in the clinical management of TB.

## 1. Introduction

Tuberculosis (TB), caused by the bacterium *Mycobacterium tuberculosis (Mtb)*, is currently the world’s deadliest infectious disease due to a bacterial infection and the second leading infectious disease killer after Covid-19. Of the 10 million new cases of TB in 2021, nearly 5% of infections were accounted for by multidrug resistant tuberculosis (MDR-TB) or extensively drug resistant (XDR) strains (Chakaya et al., 2021), with the highest burden seen in the WHO European Region including Ukraine, Moldova, Belarus and Russia. The ongoing war and humanitarian crisis in Ukraine and bordering countries is predicted to result in increased MDR-TB cases and disruption of healthcare services (Holt, 2022). Additionally, the Covid-19 pandemic has resulted in reduced access to TB diagnosis and treatment, reversing decades of progress in disease management globally. The WHO has now called for entirely new strategies to meet the goals for ‘*End TB’*, which aims to reduce TB deaths by 95% by 2035 (Chakaya et al., 2021).

While there have been significant new developments in TB diagnosis and drug discovery, the duration of TB treatment is extremely long (6-12 months for drug-susceptible TB, 12+ months for XDR) (Chakaya et al., 2021; Prasad and Gupta, 2015), often leading to treatment noncompliance. This, coupled with other factors including presence of comorbidities, patient health and socioeconomic status and increasing drug resistance make it imperative to understand early predictors of unsuccessful TB treatment outcomes to identify patients needing tailored treatment approaches, such as directly observed therapy (DOT) or extended treatment course (Mdluli et al., 2015; Zumla et al., 2015).

Predicting treatment prognosis in TB based on the relationships between different features and disease outcomes is an important facet of managing TB clinically. With the increasing availability of patient electronic health records (EHR) which provide real-world multi-domain case information, it is now feasible to build prediction models to determine important predictors and estimate an individualized probability of a specific endpoint within a defined period of time (Peetluk et al., 2021). The use of imaging techniques such as Chest X Rays (CXR) and computed tomography (CT) scans has also been shown to provide high sensitivity as a diagnostic tool and additional insight into TB disease prognosis (Huang et al., 2016; Ordonez et al., 2020). Once diagnosis of TB has been confirmed, it becomes vital that clinical healthcare workers make appropriate treatment decisions based on their own experience as well as on the individual clinical presentation of the disease. Delays in treatment initiation or providing inappropriate treatment to treat drug-resistant strains results in poor prognosis with increased clinical severity and risk of death (Lino Ferreira da Silva Barros et al., 2021).

The NIAID TB portals database is an invaluable resource for TB EHR data and is continuously updated with new patient information (Rosenthal et al., 2017). The database, as of January 2022, contains multimodal linked socioeconomic/geographic, clinical, laboratory, radiological, and genomic data from 5060 international TB patients from 10 countries with a high MDR-TB burden including Ukraine, Moldova, Georgia, India, and Belarus. With the rapid insurgence of drug resistant TB, it becomes critical to study the impact of type of resistance and infecting strain along with its genotype to better determine the course of clinical progression and treatment. Additionally, several studies have focused on the clinical and demographic data, pathogen genomics data or the imaging data modalities separately to decipher associations and make predictions (Koo et al., 2020; Peetluk et al., 2021; Rosenfeld et al., 2022). An unbiased approach that integrates all modalities of host and pathogen data available in the clinical setting is necessary to determine the most useful predictors for TB prognosis.

In this study, we analyze clinical data from drug-sensitive pulmonary TB, MDR TB and XDR TB patients across different geographical populations and implement machine learning to identify correlates of patient, drug, and pathogen features with the type of drug resistance and prognosis of treatment in individual patients. In addition, we compute drug interaction FIC scores to determine if the drug interactions among prescribed drugs play a significant role in the treatment outcome and determine the drug combinations that are most significantly associated with clinical success in drug resistant TB.

While prior studies on the TB portals data have focused largely on individual modalities, and evaluated models by cross-validation (Gabrielian et al., 2020; Sauer et al., 2018; Wollenberg et al., 2022), we conduct additional validation of our predictions on newer patient data that is populated in the TB portals database, thus providing more rigorous evaluation with new patients. We determine the most significant predictors associated with successful and unsuccessful clinical outcomes at the individual level for every patient. Overall, our integrated analysis of clinical, radiological, and genomic features aids in understanding personalized responses to TB infection and will help in clinical management of TB.

## 2. Results

The overall workflow adopted in this study to analyze EHR of TB patients is described in Figure 1. There were a total of **5060** patients in the dataset at the time of analysis, spanning 10 countries. The dataset is largely dominated by patients from Eastern Europe, particularly Moldova, Georgia, Ukraine and Belarus, which carry a high burden of drug resistant TB cases. The dataset had 203 features, which we first analyzed as 3 different modalities: a) patient socio-demographic and clinical characteristics b) radiological imaging attributes derived from chest X-Rays and CT scans and c) pathogen drug susceptibility and genomic mutations implicated in resistance to individual drugs. The dataset originally described 5 different treatment outcomes, as listed in Table 1. We pooled the outcomes ‘cured’ and ‘completed’ as they depicted a ‘successful outcome, while outcomes ‘failure’, ‘died’ and ‘palliative care’ were pooled together as ‘failure’, depicting unsuccessful outcome of treatment. After removing samples that had outcomes ‘Still on treatment’, ‘unknown’ or ‘not reported’, there were a total of 4139 TB patients with these two outcomes of success and failure that were then considered for further analysis for each modality.

**Figure 1.**
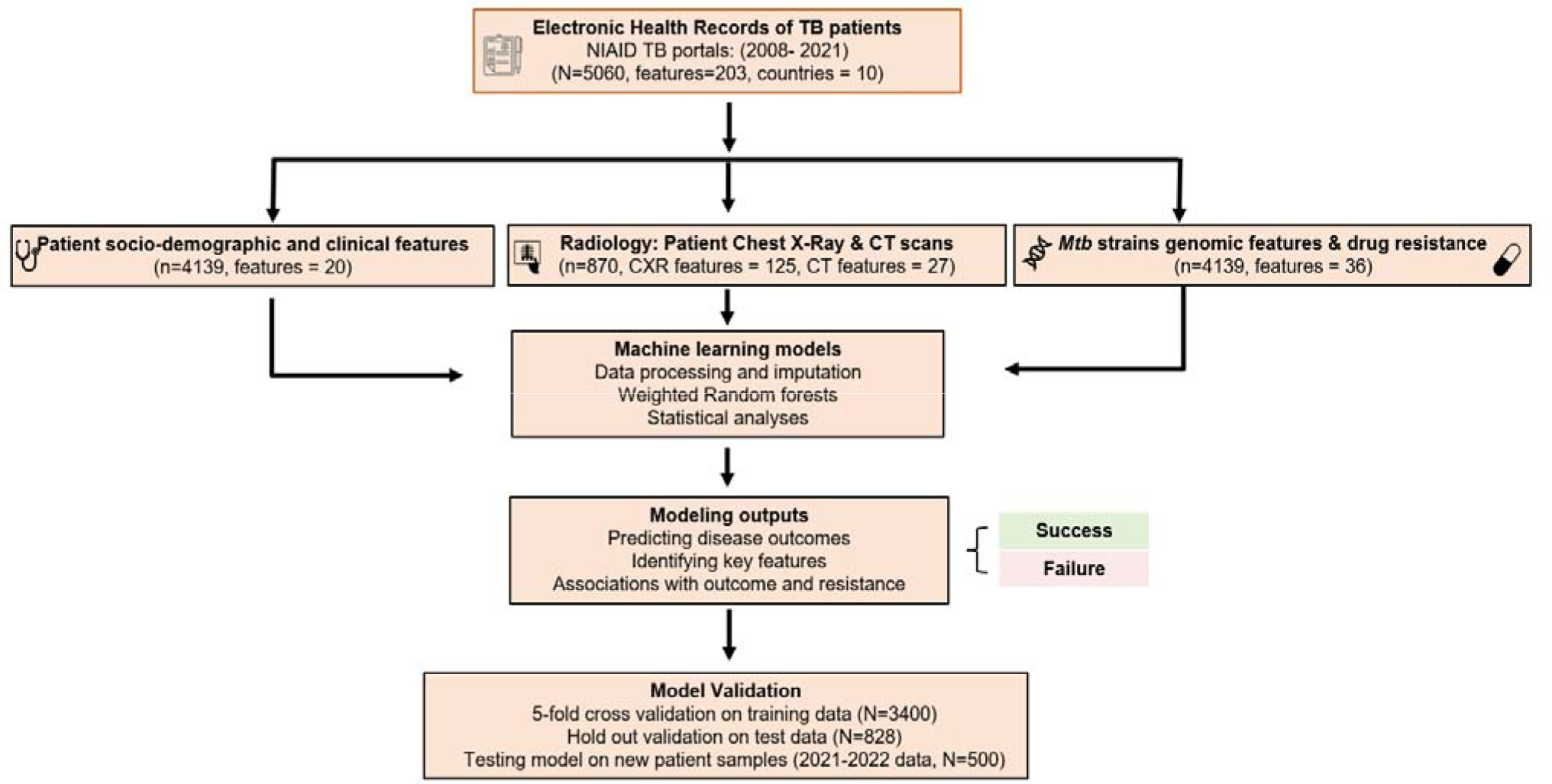
Workflow adopted in this study to analyze Electronic Health records of TB patients. Electronic health records for tuberculosis patients were retrieved from the NIH TB portals database. A total of 5060 patients from 10 different countries were considered for the analysis, with over 200 features available representing pathogen genomic features, patient clinical and social features as well as radiological features derived from patient chest X rays. We analyzed each of these categories separately. Data analysis involved building machine learning models (Random forests) and conducting statistical analyses to predict disease outcomes – grouped as Success and Failure, respectively. Feature importance was performed using Shapley analysis as well as hypergeometric tests to determine the predictors associated with both success and failure. A final unified model was built comprising top features across all modalities. All model performances were evaluated by 2 types of validation a) Cross-fold validation (k=5) where the input data was split into training and test sets b) Predicting outcomes on newer patients that were populated in the TB portals database.

**Table 1.** Definition of treatment outcomes considered for our study

| Outcome | Definition | # of patients | Outcome assigned for this study |
| --- | --- | --- | --- |
| Cured | Treatment completed as recommended by the national policy without evidence of failure AND three or more consecutive cultures taken at least 30 days apart are negative after the intensive phase | 2556 | Success |
| Completed | Treatment completed as recommended by the national policy without evidence of failure BUT no record that three or more consecutive cultures taken at least 30 days apart are negative after the intensive phase | 537 | Success |
| Failure | Treatment terminated or need for permanent regimen change of at least two anti - TB drugs because of: a lack of conversion by the end of the intensive phase, or a bacteriological reversion in the continuation phase after conversion to negative, or a evidence of additional acquired resistance to fluoroquinolones or second - line injectable drugs, or a adverse drug reactions (ADRs) | 481 | Failure |
| Died | A patient who dies for any reason during the course of treatment | 565 | Failure |
| Palliative Care | An approach that improves the quality of life of patients and their families facing the problems associated with life-threatening illness, through the prevention and relief of suffering by means of early identification and impeccable assessment and treatment of pain and other problems, physical, psychosocial, and spiritual | 101 | Failure |

### 2.1 Statistical exploration of TB Portals provides insights into shared host and pathogen features across populations

The TB portals database had clinical data from 5060 patients collected across 10 countries spanning Eastern Europe, Asia and Africa at the time of analysis as seen in Figure 2a and is constantly being populated with new patients from additional countries. Data from Moldova, Georgia and Belarus are most prevalent, followed by Ukraine, Azerbaijan, Romania and Kazakhstan (Figure 2b). These represent a high burden of drug resistant TB cases, even though China and India have an overall higher burden of TB (Chakaya et al., 2021). As described in the methods, we pooled all outcomes into either Success or Failure, with Successful outcomes seen to be present at least 3-fold more than Failure (Figure 2c). We take this data imbalance into consideration while building and analyzing our models. We also observe a higher male population with TB compared to the female population both overall (Figure 2d), as is consistent with most TB reports globally. Majority of the TB cases in the dataset are New or first instances of TB reported for that patient, while more than 500 patients show instances of relapse, as per the case definition (Figure 2f) The distribution of BMI (Figure 2e) shows a lower average BMI in patients with failed treatment outcomes (average BMI 19.2) compared to those with successful treatment outcome (average BMI 21.4).

**Figure 2.**
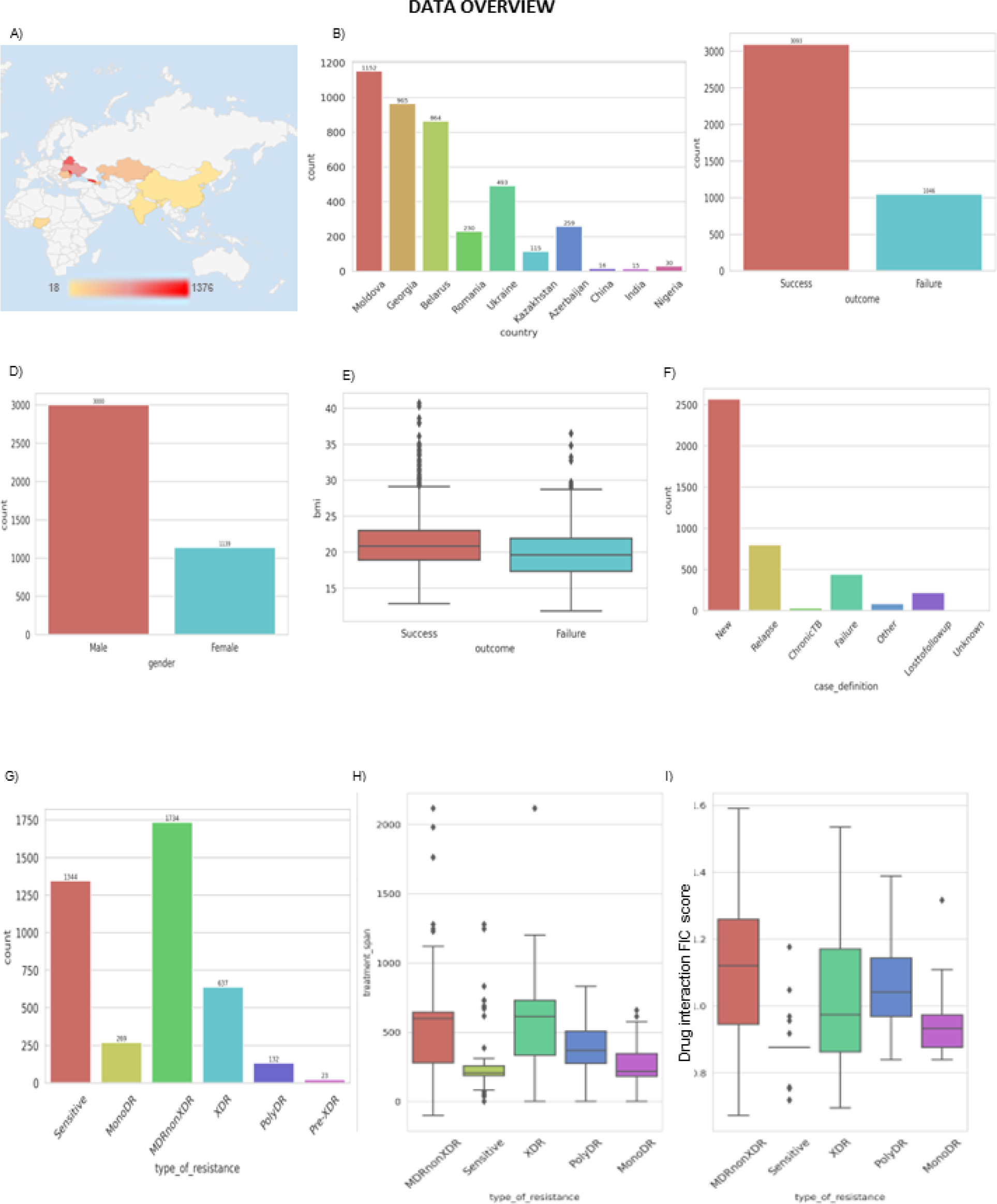
Overview of the EHR data present in the TB portals database. a) Geographical distribution of patients in the database shows countries in Europe, Asia and Africa represented in the dataset. Distributions of b) number of patients per country for all 10 countries present in the database c) 2 pooled outcomes of Success and Failure d) patient numbers by gender and e) different numbers of patients by case definition. e) Distribution of BMI levels seen across outcomes of success and failure. Panels g) - i): g) Drug susceptibility in *Mtb* strains isolated from patients implicated in resistance to individual drugs, h)the distribution and average treatment times for each type of resistant strain, and i) the drug interaction FIC scores for treatment regimens to treat different kinds of resistant Mtb as calculated by INDIGO-MTB.

There are 27 different Mtb families represented in the data encompassing 3 major genetic lineages L2, L4 and L1, with the L2–Beijing sub-lineage seen to be most prevalent in patients across countries. L2 has received much attention due to its high virulence, fast disease progression, and association with antibiotic resistance (Gagneux, 2012). The *Mtb* strains show varying drug susceptibilities to the 28 different drugs in the clinical dataset (Fig 2g). There are 6 types of drug susceptibilities observed in the Mtb strains namely a) Sensitive, implying no resistance to any anti-TB drugs b) Mono-DR, where resistance is seen to one first-line anti-TB drug c) Poly-DR, where resistance is seen to more than one first line anti-TB drug, d) MDR-non-XDR, where resistance is seen to at least both isoniazid and rifampin, e) Pre-XDR: TB caused by Mtb strains that are Multidrug resistant and rifampicin resistant (MDR/RR TB) and also resistant to any fluoroquinolone and f) XDR-TB caused by Mtb that is resistant to isoniazid and rifampin, plus any fluoroquinolone and at least one of three injectable second-line drugs (i.e., amikacin, kanamycin, or capreomycin)(Chakaya et al., 2021; Johnson et al., 2006; Mitchison, 2005). The MDR-non-XDR type of resistance is observed to be most prevalent across infected populations in the dataset. Patients with drug resistant TB show a corresponding increase in treatment times compared to drug-sensitive TB, with the longest average treatment duration seen for patients with MDR and XDR TB (Figure 2h).

Treatment regimens vary based on the type of resistance being treated. For the different drug regimens assigned per patient (758 unique regimens in total), drug-interaction scores were calculated using the machine learning tool INDIGO-MTB (Ma et al., 2019). These scores provide a quantitative description of the nature of interaction between drugs (synergy, additivity, and antagonism), with lower scores associated with strong synergy among the drugs in each regimen. We observe that regimens used to treat drug sensitive TB have the strongest synergy and correspondingly lower treatment times (figures 2h, 2i), while drug combinations used to treat resistant TB cases show a wider range including weak synergistic and antagonistic interactions. These interactions also lead to increased treatment times for drug resistant TB, as seen with MDR-TB and XDR-TB duration relative to drug sensitive TB.

### 2.2 Modeling socio-demographic and clinical modalities reveal comorbidities most predictive of treatment failure

We grouped different features by modality and built models separately for each modality as illustrated in Figure 1. There were a total of 20 features describing patient clinical, social and demographic aspects. After cleaning and imputation of the data, we built a random forest machine learning model as discussed in the Methods, and conducted 5-fold cross-validation, and hold-out validation on blinded test data as well as on newer patient data populated in the TB portals database from August 2021-January 2022. Our model predictions had accuracies of 78.9%, 84.7% and 80.7% as well as Matthews correlation coefficient (MCC) values ranging from 0.4 to 0.47 for the cross validation, hold out validation, and validation on new patient data sets respectively (Figure 3A - E). The MCC is a robust statistical metric which produces a high score only if the predictions have high precision, recall and accuracy (i.e. all of the confusion matrix categories) and accounts for the biased distribution of Success and Failure in the dataset.

**Figure 3.**
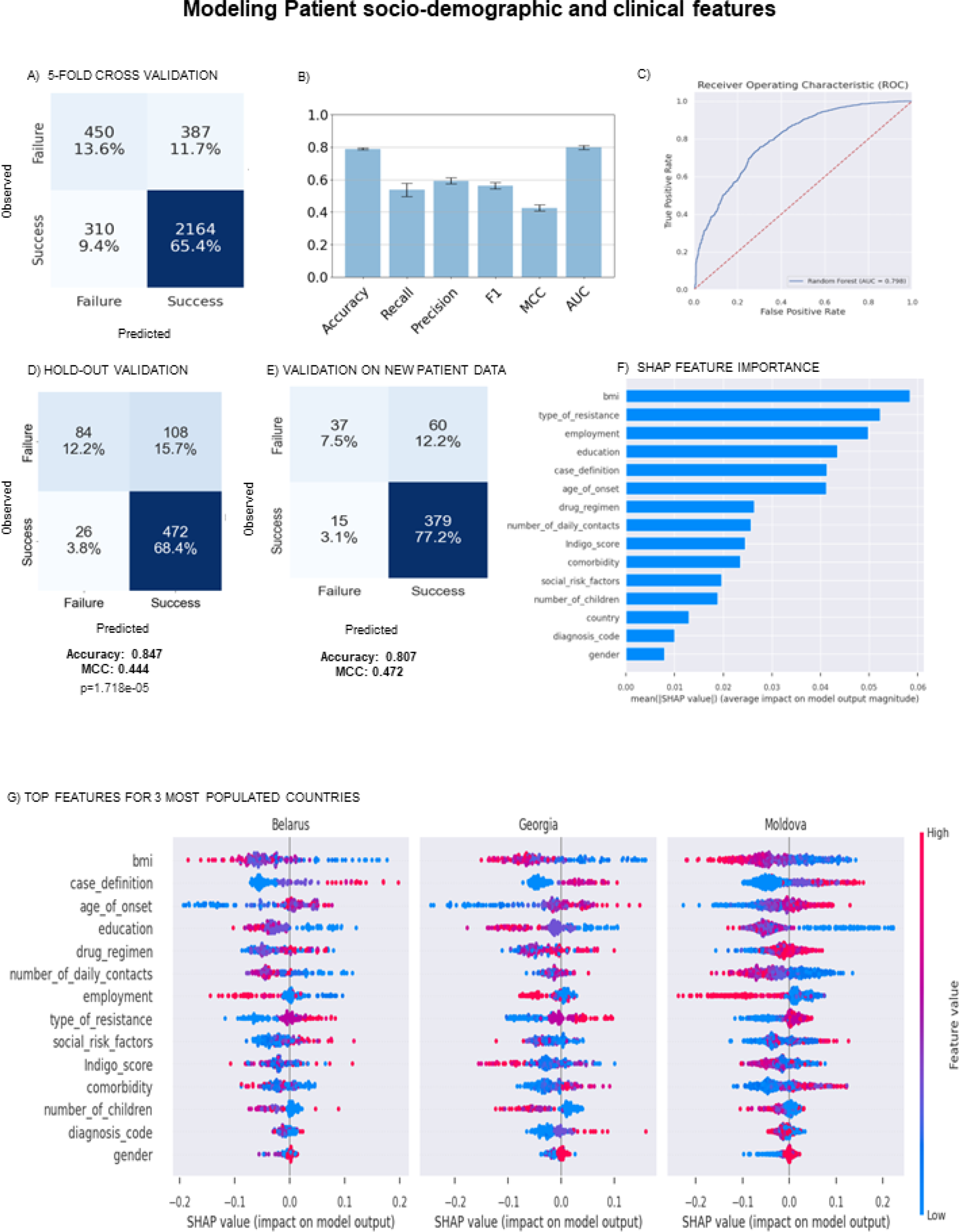
Modeling patient socio-demographic and clinical features. a) Model predictions on 80% training data using 5-fold cross validation b) model evaluation metrics of Accuracy, Recall, Precision, F1 score, MCC, AUC values and Correlations c) ROC curve generated on the training data with 5-fold cross validation d) Model predictions on 20% hold-out validation data e) model predictions on newer patient data f) top features predictive of treatment outcome g) top features and their shapley values across the 3 most populated countries in TB Portals. Each dot represents a single patient, with blue color representing lower values for each feature. The X axis represents the impact of the feature value driving model outcomes, with values above 0 associated with failure, and those below 0 associated with success.

The most important predictors of Failure were determined by Shapley analysis, a game theoretic approach that evaluates the impact of removing each feature on model accuracy (Merrick and Taly, 2020; Winter, 2002) (Figure 3F). The top predictors were BMI, type of resistance observed, education, employment, case definition, drug regimen used and its corresponding Drug interaction FIC scores, comorbidities, social risk factors and gender. To further inspect what values among these features were truly associated with failure, we analyzed these feature value distributions for patients for each country as well as across countries. Figure 3G shows the shapley feature value distributions for the 3 most populated countries in the dataset, namely Belarus, Moldova and Russia. We observe similar trends in the values for top features across all countries (including Ukraine, Azerbaijan, Romania - Supplementary Figure 1). BMI values rank as the highest predictor, with lower values (in blue) associated more with failure compared to higher values. Higher age of onset (red), higher resistance (red/purple), lower education (blue) are significantly associated with failure.

We delved further into each feature category to determine associations between multiple feature levels and failure, with the significance determined by hypergeometric cumulative distribution testing (Methods). Patients with BMI values less than 18, classified as *“Underweight”* have strong associations with treatment failure (p=1.32 × 10-23), compared to those that were classified *healthy, overweight* or *obese*. (Fig 4). We also observe more failure in patients with disability (p= 3.55 × 10-25), lower employment (p=2.09 × 10-05) and lower education levels (p=1.26 × 10-25). Prior history of alcohol (p=4.46 × 10-34), drug abuse (p=1.01 × 10-16) and smoking (p=3.76 × 10-10) impact the treatment outcome as well. Patients with comorbidities HIV (p=3.25 × 10-32), Anemia (p=2.34 × 10-31) or Hepatitis B/C (2.6 × 10-04) are also seen to have poorer outcomes. While Diabetes is a prevalent comorbidity commonly seen in TB patients, it is not significantly associated with poor treatment outcomes.TB is typically more prevalent in the undernourished population who are often living below poverty level and do not have access to good nutrition(Chakaya et al., 2021; Papathakis and Piwoz). Our findings are thus consistent with these reports, highlighting the importance of nutrition and income to have successful treatment outcomes. Anemia in tuberculosis is most often due to nutritional deficiency or malabsorption syndromes, correlating with lower BMI. TB associated anemia has also been linked to distinct inflammatory profiles persisting after therapy as is considered a biomarker for disease severity as well (Barzegari et al., 2019; Lee et al., 2006). As observed globally, patients in this dataset also show poorer treatment outcomes in case of drug resistant TB compared to sensitive TB, with XDR and MDR TB patients faring the worst in terms of outcome.

**Figure 4.**
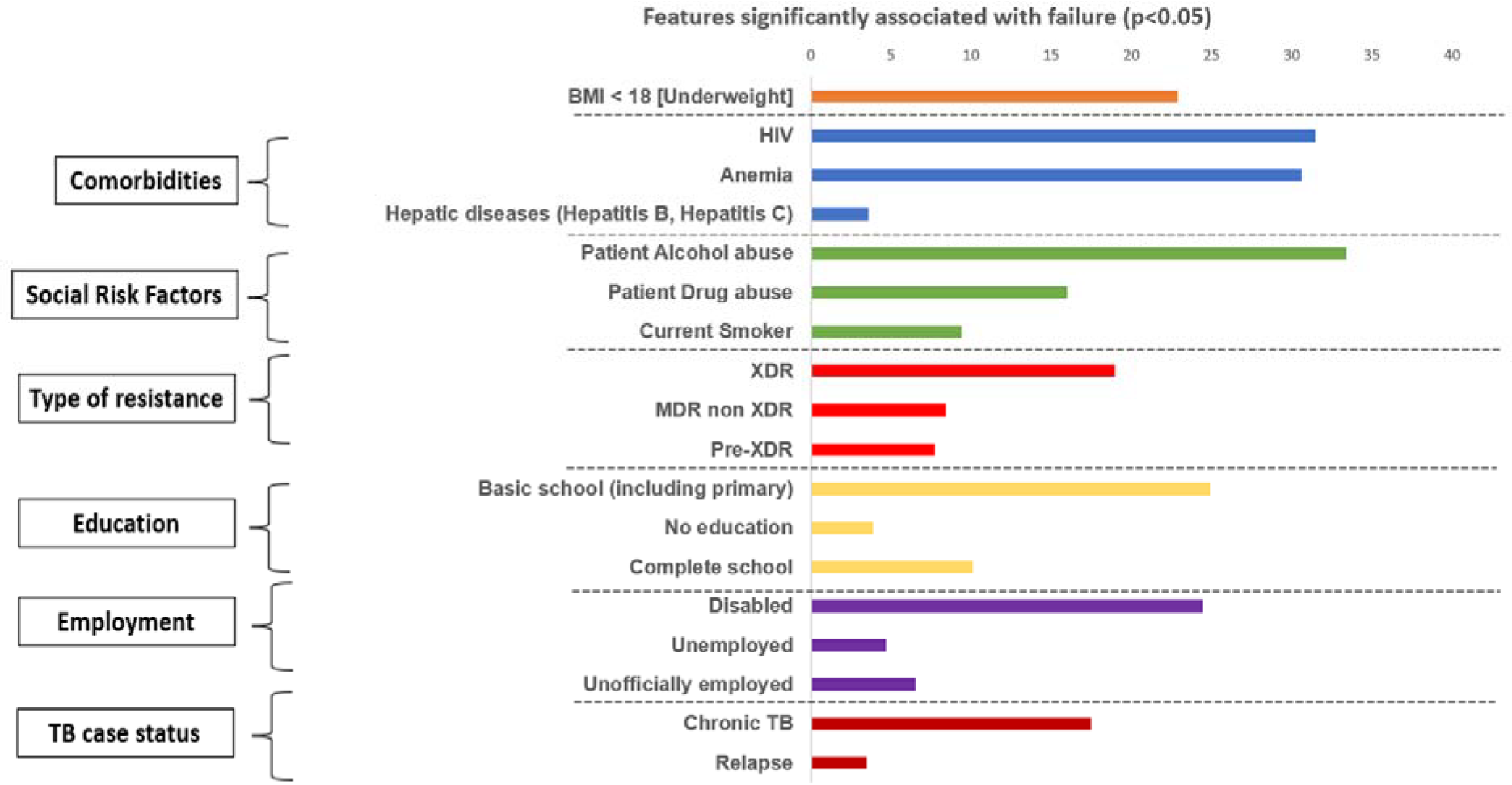
Breakdown of socio-demographic and clinical features significantly associated with treatment failure (p<0.05). Each top feature category as determined in figure 3F is broken down to depict significant associations between feature subtypes and failure, highlighted by different colors. All pvalues are significant (p<0.05) after multiple hypothesis correction (fdr <0.1). The scale represents negative log p-values determined by hypergeometric tests.

### 2.3 Analysis of pathogen features identifies mutations linked with treatment failure and drug regimens associated with successful treatment of drug resistant TB

Across all populations, the data reports a total of 27 different clinical *Mtb* strains belonging to 5 different *Mtb* families with varying drug susceptibilities (Supplementary table 1). The Beijing, H3 and T1 families were most prevalent in all infections observed, and strains from these families show all types of drug susceptibilities, from sensitive TB to XDR TB. Our modeling analysis using the pathogen genomics and drug resistance modality could predict treatment failure with accuracies of 69%, 72% and 74% using cross-validation, hold-out validation and the new patient data respectively (fig 5a-d). We observe that the prediction accuracy was relatively lower using pathogen features alone, compared to using clinical features. Feature analysis revealed that higher numbers of *Mtb* colonies as determined by culture and the presence of mycobacterial growth were strongly associated with treatment failure. Of the 28 different drugs present in treatment combinations across all regimens, resistance to rifampin, isoniazid, kanamycin, streptomycin, ethambutol, capreomycin and amikacin were more significantly predictive of treatment failure as determined by shapley feature analysis (fig 5e). We analyzed the SNPs associated with each strain as reported, and identified mutations in genes gyrA, rpsL, and katG to be strongly associated with failure. Mutations in the gyrA gene, particularly at positions 90, 91 and 94 have been frequently reported among fluoroquinolone resistant Mtb (FQr-MTB) isolates.

**Figure 5.**
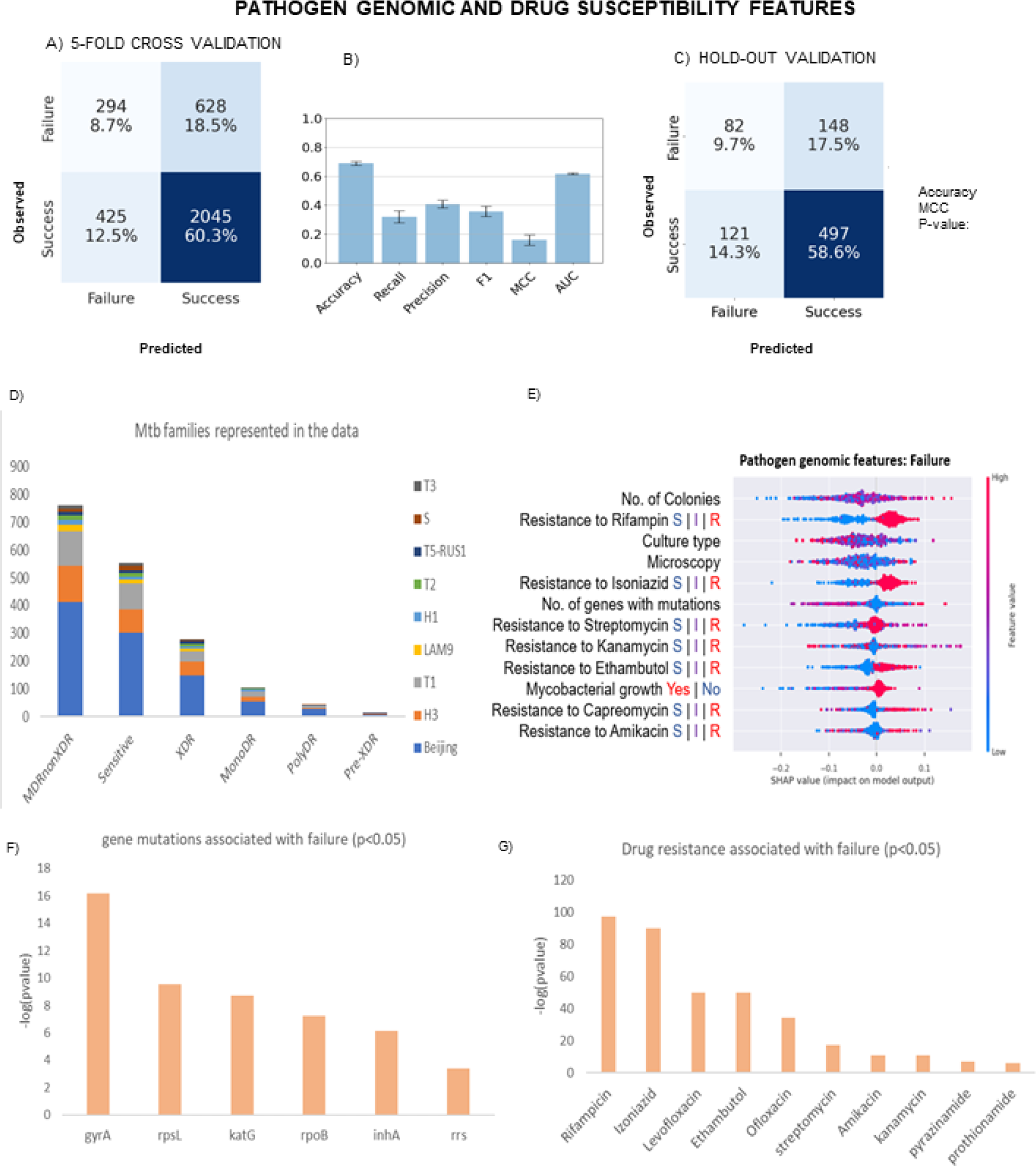
Modeling pathogen genomic features and drug susceptibilities. a) Model predictions on 80% training data using 5-fold cross validation b) model evaluation metrics of Accuracy, Recall, Precision, F1 score, MCC, AUC values and Correlations c) Model predictions on 20% hold-out validation data d) Top features associated with failure e) *Mtb* families represented in the data with varying drug sensitivities. f) *Mtb* genes where mutations are significantly associated with poor treatment outcomes (p<0.05) and g) *Mtb* resistance to drugs significantly associated with failure (p<0.05).

We analyzed the drug interactions among treatment regimens used to treat TB caused by different Mtb strains of varying drug susceptibilities. The standard 4-drug regimen of HRZE was synergistic (FIC score 0.8) and associated with treatment success of sensitive and mono-DR TB cases. Among MDR and XDR TB cases with high rates of failure, we conducted a hypergeometric cumulative distribution test to identify regimens most associated with success in these instances. Treatment regimens involving combinations of Bedaquiline, Levofloxacin, Clofazimine and Amoxicillin see more success in treating MDR and XDR TB (p = 3.25e-05). This is concordant with recent reports on new regimens approved by the FDA involving Bedaquiline and Levofloxacin (Burki, 2019). Drug interaction FIC scores for all regimens are highlighted in Supplementary Table 2.

### 2.4 Lung volume, pleural effusion, and bronchial obstruction are significantly predictive of treatment failure and associated with drug resistant TB

Radiological imaging using CXR and/or CT scans are typically used to aid clinicians in reaching a diagnosis of TB and monitoring clearance of infection. They complement *Mtb* culturing and symptoms. Imaging can reveal TB lesions of differing size, shapes and characteristics (eg. cavitation) occurring anywhere in the lungs. This dataset had 406 patients with at least 1 X-ray available, with an average of 3 X-rays per patient taken over their course of treatment. We chose the CXR taken closest to the date of treatment initiation to assess if there were any features that would help indicate treatment prognosis at that time. We validated our model on 289 new patients with CXR available in the January 2022 dataset. As only 59 additional patients had CT scans available, we conducted validation with new patient entries from TB Portals using CXR data alone.

Models built on CXR and CT data individually could predict treatment prognosis reasonably well (accuracies of 74% and 83% respectively) (Figure 6 a-f). Validation with new patient CXR data showed an accuracy of 77% and MCC of 0.4 (Supplementary figure 2). Despite predicting outcomes with an accuracy of 83%, the CT predictions alone had poorer MCC (0.2). As a result, we pooled in the imaging features for patients who had both CXR and CT records and then conducted feature selection. The most significant TB-related manifestations that are predictive of both treatment outcome and resistance are shown in Figure 5g and Table 2.

**Figure 6.**
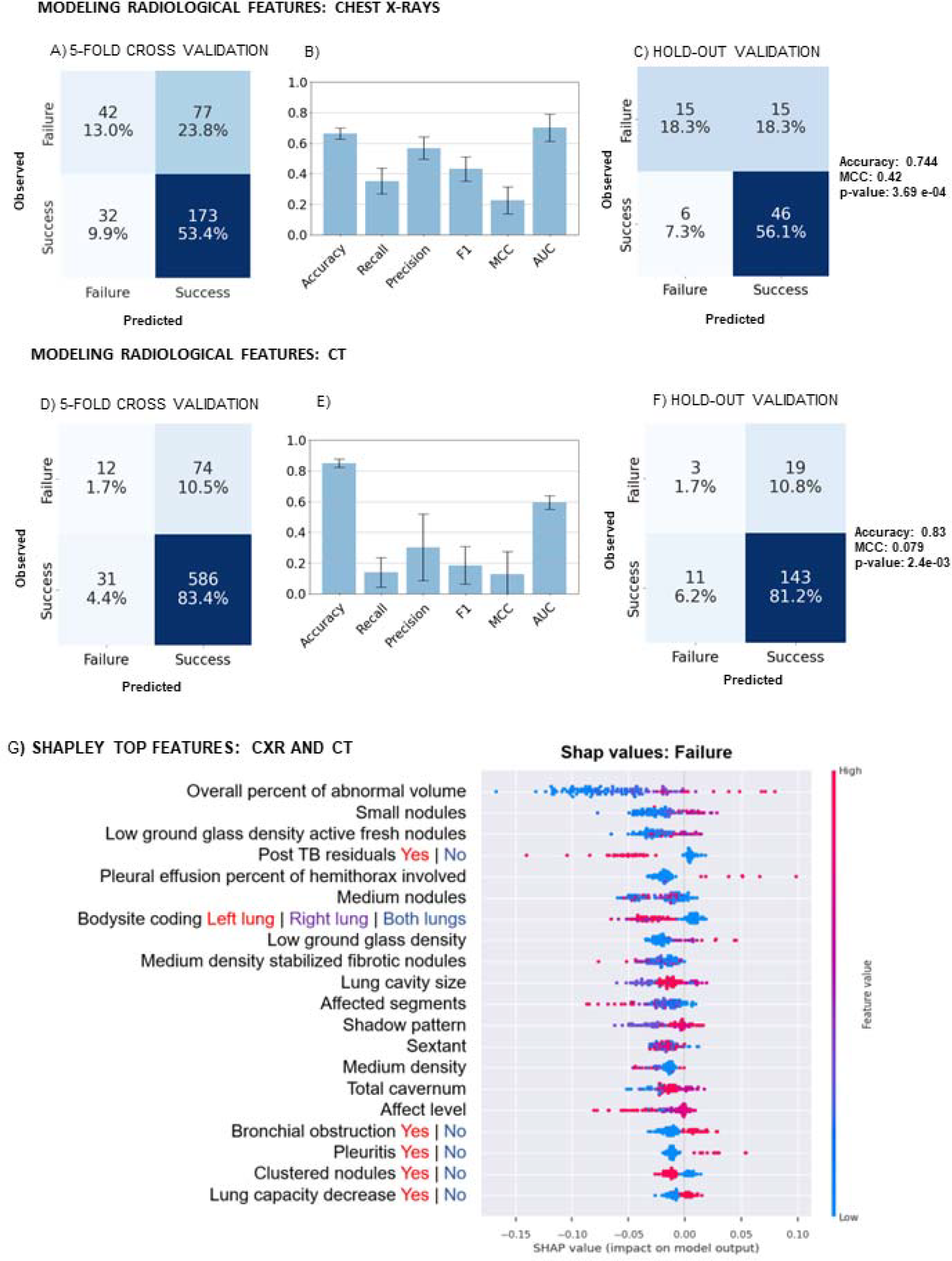
Modeling Imaging modalities (CXR and CT data). a) -c) CXR data. Model predictions on 80% training data using 5-fold cross validation b) model evaluation metrics of Accuracy, Recall, Precision, F1 score, MCC, AUC values and Correlations c) Model predictions on 20% hold-out validation data d)-f) CT data. d) Model predictions on 80% training data using 5-fold cross validation e) model evaluation metrics of Accuracy, Recall, Precision, F1 score, MCC, AUC values and Correlations f) Model predictions on 20% hold-out validation data f) Top features identified by Shapley analysis for both CXR and CT data models.

**Table 2.** Imaging features associated with treatment failure and drug resistant TB

| Chest Imaging Feature | Association with Failure<br>($p < 0.05$) | Association with Resistance<br>($p < 0.05$) |
| --- | --- | --- |
| Bodysite: Both lungs | 4.16E-06 | 9.09E-04 |
| Pleuritis: Yes | 4.15E-05 | 2.11E-60 |
| Bronchial Obstruction: Yes | 7.33E-04 | 0.04 |
| Lung capacity decrease: Yes | 0.0017 | 4.35E-04 |
| Opacity due to nodule, node and infiltrate | 0.0068 | 3.22E-04 |
| Overall percent of abnormal volume | 3.10E-13 | $p > 0.05$ |
| Small cavities: UL \| UR | 4.30E-11 | $p > 0.05$ |
| Small cavities: ML \| MR | 5.40E-06 | $p > 0.05$ |
| Collapse: UL \| UR | 7.50E-05 | $p > 0.05$ |
| Enlarged lymph nodes: Yes | 4.75E-04 | $p > 0.05$ |
| Nodes more than 10mm | $p > 0.05$ | 7.16E-11 |
| Affect Pleura: Yes | $p > 0.05$ | 0.022 |
| Post TB residuals: Yes | $p > 0.05$ | 0.0028 |
| LL \| LR calcified or partially calcified nodules: Yes | $p > 0.05$ | 6.03E-05 |
| ML \| MR calcified or partially calcified nodules: Yes | $p > 0.05$ | 3.12E-08 |
| UL \| UR calcified or partially calcified nodules: Yes | $p > 0.05$ | 7.00E-11 |

The TB portals database annotates each CXR into sextants to highlight regions that were afflicted in the patient’s lungs viz. Upper Left (UL), Upper Right (UR), Middle Left (ML), Middle Right (MR), Lower Left (LL) and Lower Right (LR). TB is typically manifested as an upper respiratory infection, but it has been shown to infect different parts of the lung based on disease severity(Koo et al., 2020). Our analysis indicated that the overall affected abnormal volume across both lungs was more significant in predicting poor disease prognosis rather than individual affected sextants. The presence of bronchial obstruction, pleuritis (inflammation of the tissues that line the lungs and chest cavity), a decrease in lung capacity and the presence of lung opacities due to nodules, nodes (seen in shadow patterns) and airspace disease (infiltrates) were strongly predictive of both treatment failure and were significantly associated with drug resistant TB. It is important to note that we analyzed the clinical presentation of drug resistance as seen through imaging data for our study, as drug resistance can only be predicted directly by culturing the infecting *Mtb* strain. Interestingly, the presence of calcified or partially calcified nodules across most sextants, nodes larger than 10mm, and the presence of post TB residuals were seen to be associated with DR-TB cases, but they were not especially predictive of treatment failure (Table 2). The overall percentage of abnormal lung volume, which is a quantification of TB severity, was most predictive of treatment failure. Similarly, lymph node enlargement (lymphadenopathy)(Ahmed et al., 2011; Rosenfeld et al., 2022), and collapse of the Upper or Middle lung were indicative of a poor treatment prognosis. Chest imaging usually reports several hundred features (152 imaging features in this dataset), and our analysis identified 21 TB manifestations to be most clinically significant, which will thus help clinicians and radiologists make more informed decisions about treating individual patients when making TB diagnosis from patients’ CXRs and CTs.

### 2.5 Integrated multi-modal analysis outperforms model predictions of individual modalities

Modality-wise analysis helped identify the important features in the context of treatment prognosis for drug sensitive and drug resistant TB. The original dataset has 203 features across all modalities, not all of which are clinically relevant. Our analysis on individual modalities revealed the most predictive features per modality (15 top socio-demographic features, 23 top pathogen genomic features and 21 top imaging features), resulting in 59 top features across all categories.

We next analyzed correlations among the final top features across modalities (Figure 7c and Supplementary Table 3). We find high correlations among features within each modality, as well as between features across modalities. The type of resistance is significantly associated with the drug regimen and corresponding drug interaction FIC score. Drug resistance was also correlated with the imaging attributes of increased bronchial obstruction and affected lung segments as seen in the clinical presentation of drug resistant TB. Disease severity as reported by diagnosis code correlated with the number of *Mtb* colonies and the affected lung segments. Social risk factors and comorbidities show associations with gender and education; employment and BMI are correlated, highlighting the socio-economic impact in TB prognosis and management.

**Figure 7.**
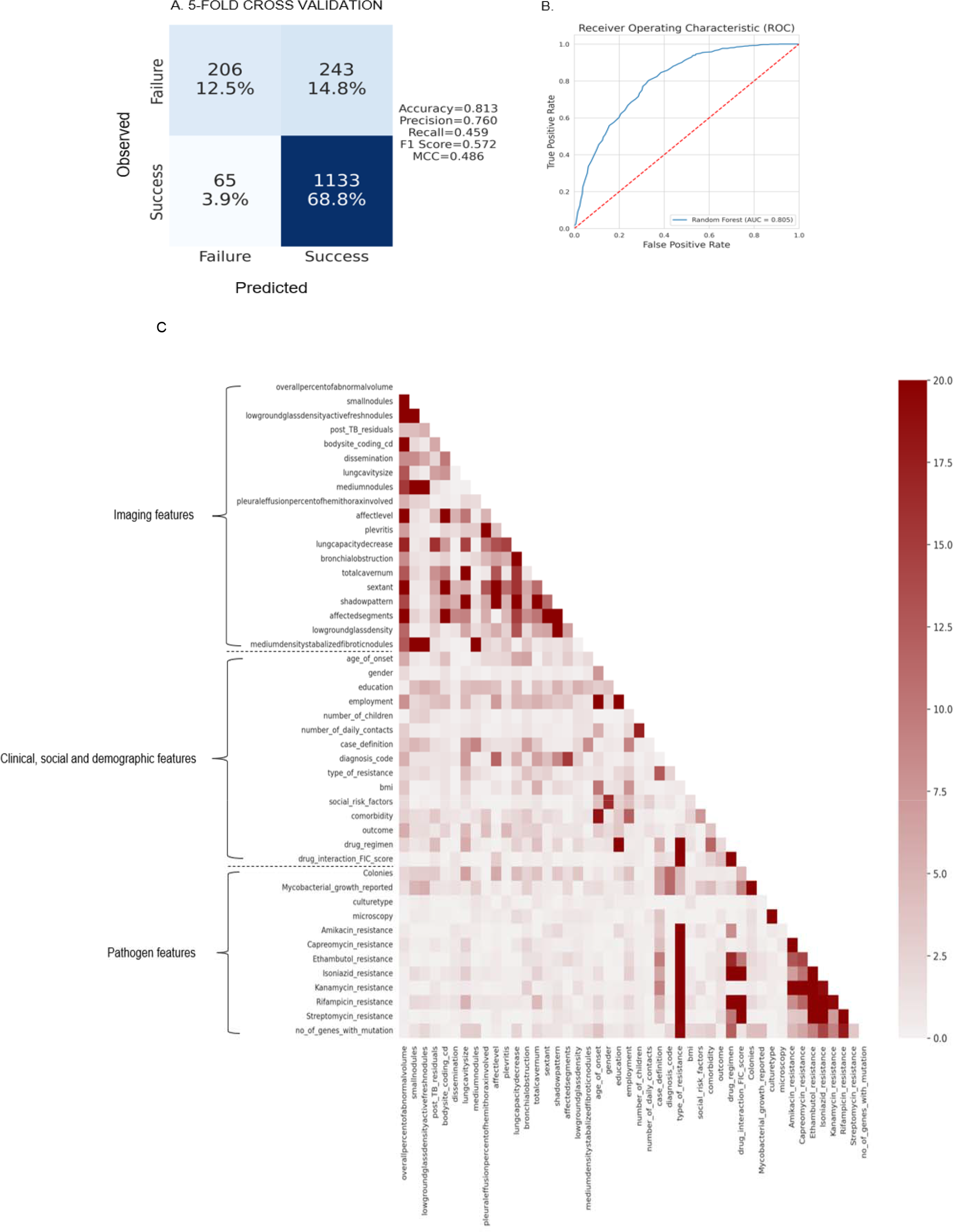
Feature associations between top features across all modalities. The color scale is based on -log(p values) for correlations between these features.

A multi-modal machine learning model with these top features across modalities could predict treatment failure with an accuracy of 81.3% and an AUC of 0.805 and MCC of 0.486 (Figure 7a and 7b). For comparison, the AUC values were 0.798, 0.59 and 0.65 and the MCC values were 0.41, 0.19 and 0.22 for clinical, pathogen and imaging models separately. These results are comparable or better than prior models in literature that predict TB treatment failure (AUC = 0.70 (Kalhori et al.), 0.74 (Sauer et al., 2018) and 0.79 (Koo et al., 2020)). These results are especially encouraging, as EHR data was collected across several hospitals within each country, each of which had their own data collection and reporting criteria. As a result, and as is common with real-world patient data, there were several redundancies, missing information and poor labeling, which made the data noisy. Our final integrated model is thus quite robust and can predict treatment outcomes with over 81% accuracy despite these data limitations. Our model can also handle missing values effectively. We also built a machine learning model with only samples that had no missing data (without imputing missing values) which yielded a similar accuracy of 80.3%. The final model developed in our study can thus guide the development of machine learning models to predict TB prognosis and determine optimal treatment regimens based on real world multi-modal data.

## 3. Discussion

Tuberculosis remains a significant challenge globally, but especially now in the aftermath of the Covid-19 pandemic (Chakaya et al., 2021; Zhou et al., 2020). There were ∼1.5 million deaths reported due to TB in 2020, marking a significant increase in TB mortality. The Covid-19 pandemic has had devastating effects on every aspect of global health, but TB services have been disproportionately affected (Pai et al., 2022) with reduced access to care and treatment. Of particular concern are the regions in Eastern Europe which have a high incidence of TB, TB-HIV co-infection, and multidrug-resistant TB (MDR-TB) in the region (Chakaya et al., 2021) (Marais et al., 2013). The ongoing war and humanitarian crisis in Ukraine has affected the care of patients and the efficiency of healthcare systems in the area, including both Eastern and non-Eastern European countries. Given these scenarios, it becomes especially important to analyze available clinical data to aid in TB prognosis and guide optimal treatment decisions for each patient (Falzon et al., 2018; Gröschel et al., 2018; Lange et al., 2020; Lino Ferreira da Silva Barros et al., 2021; Olaru et al., 2016)

The increasing availability of patient EHR comprising multi-modal data can enable precision and personalized medicine initiatives to better understand TB and other diseases (Piekos et al., 2022). We analyzed EHR data from over 5000 patients across 10 countries from the NIH NIAID TB portals. We implemented a ‘transparent’ machine learning model to identify patient, drug, and pathogen features predictive of drug resistance and treatment prognosis in individual patients. Our analysis of over 200 different features across different host and pathogen modalities revealed a smaller set of 59 significant predictors associated with successful clinical outcomes at the individual level. We also determined significant associations between different feature subtypes and treatment failure, as well as correlations within these top features across different modalities. In addition, we identified drug combinations that are associated with clinical success in drug resistant TB. Finally, in contrast to prior predictive models that were evaluated using only cross-validation, we conducted additional validation of our predictions on newer unseen patient data that is populated in the TB portals database, which provides more rigorous evaluation. We also observe that features related to nutrition, particularly lower BMI and the presence of HIV and Anemia are significantly associated with failure, which we expect to worsen with conditions of war in these regions.

There are some limitations to our study. As with any real-world patient data, the TB portals dataset has several missing and noisy information, since it collects information from multiple hospitals across 10 countries, each with their own collection and reporting protocol. Further, there is a significant imbalance in the outcomes, with Success more prevalent than Failure. We address these limitations by considering Random forests with an inverse weighting approach, with Failure assigned higher weight than Success in order to account for such imbalance. Random forests perform better than other modeling approaches as they are able to work with mixed data types and with missing values (Peetluk et al., 2021). Despite the missing information, our models were robust and performed significantly well and can be used to assess clinical outcomes and resistance. Treatment regimens provided in the TB portals are not reported sequentially for each patient over time, but rather are listed as a single instance collectively. As a result, some patients have treatment regimens of more than 10 drugs, which is typically not the case. Our drug interaction FIC scores determined by INDIGO-MTB are computed with the assumption that these drug regimens are provided simultaneously. It would be more clinically relevant to calculate sequential drug interaction scores based on when they are provided for each patient longitudinally (Chung and Chandrasekaran, 2021; Cicchese et al., 2021). Such sequential treatment information should be made available in the EHR databases. Other limitations include that genomic information linked with clinical cases is only available for a few genomes, based on sequencing availability at the time of collection. Whole genome sequencing is expensive in high burden TB countries and may not always be feasible. Finally, we used a transparent and mechanistic AI approach here to enable both interpretation and prediction of TB treatment outcomes. The use of black box methods like deep neural networks may lead to models with higher predictive accuracy. Nevertheless, our multi-modal mechanistic AI model’s performance is better than prior AI models in literature (Kalhori et al.) (Sauer et al., 2018) (Koo et al., 2020).

In sum, our study analyzes multi-domain information from patients across geographical regions and focuses on the urgent need to improve TB clinical management particularly in the face of increasing drug resistance. The findings of our study are especially important given various humanitarian crises worldwide in order to meet the WHO’s goals to ‘End TB’ by 2035. Our interdisciplinary study can help shift from the current one-size-fits-all approach for TB treatment to a personalized approach specific to each patient and which accounts for drug susceptibility profiles of the infecting *Mtb* strain. This study can serve as a framework for managing other drug resistant infections as well. Machine learning and Artificial intelligence based methods have shown promise in leveraging EHR data to develop personalized prediction models for addressing several infectious diseases, diabetes, cancers and other diseases to support clinical care (Bartelink et al., 2017; Tarumi et al., 2022). Our robust analysis on modeling real world clinical data from TB patients further underscores the importance of these modeling methods to assist in clinical decision making.

## 5. Methods

### Data mining

Patient EHR data was obtained from the TB portals database for patient cases available from 2008 until August 2021 [TB_portals_Update_Aug2021] after signing a data-sharing agreement. The patient data is encoded with unique IDs determined by the database, with no disclosure of the individual patient name. The database includes patients from 10 countries spanning Eastern Europe, Central Asia, and Africa with a heavy burden of drug-resistant TB. It continues to be actively populated with new patient data, including those who are still undergoing treatment. For every patient, we obtained associated information pertaining to Patient cases (clinical features), Radiological information (chest xray images, CT scans and their annotations), drug regimens, biochemistry, drug sensitivity profile for the pathogen associated with each infection and the corresponding specimen. The TB portals data contain de-identified multi-domain TB patient case data. They utilize a uniform data dictionary with generally accepted medical terminology and data field values. The data collected comes from a range of sources – clinical trials, research studies, as well as routine collection of atypical patient cases receiving medical care. There is no single identifiable data collection protocol that is uniformly enforced. Therefore, TB Portals data are structured and should be regarded as a natural history study, not an epidemiological study.

### Data processing

The dataset had missing values for several features as well as entries with values ‘Not Reported’, ‘Unknown’, ‘Others’, ‘Not Specified’. These were collectively labeled as “NaN” for further processing. Duplicate patient records as determined by patient id numbers were removed. Entries with conflicting values (eg. entries reporting ‘Yes AND No’ or ‘Resistant And Sensitive’) were also reported as NaN for that feature if the entries for other features for that patient were not conflicting. For every patient, there were 203 variables of mixed data types, with 11 numerical and 192 categorical features. The categorical features were encoded into numerical values, with numerical values assigned based upon the number of levels an attribute, starting from 0. For example, the attribute ‘education’ has six levels, ranging from “no education” - which is assigned 0, to “college and higher” which is encoded as 6. This process is repeated for all categorical attributes accordingly.

### Culture and Microscopy

For culture types, the first culture report of the number of colonies identified in the specimen were considered. Instances which don’t report the number of colonies were mapped to the individual sample results associated with specimen id. Growth of mycobacteria is also reported as either positive, negative or both, as well as reports of non-specific mycobacteria and Mycobacteria other than tuberculosis (MOTT). Microscopy results describe the number of acid resistant bacteria in different fields of view, and were mapped to the following codes. For entries with multiple codes per cell, only the first entry was considered for analysis.

### Drug Sensitivity Test (DST)

The dataset describes DST results for 24 different drugs. Entries are marked R (resistant), S(sensitive) or I (intermediate) to describe observed DST profiles for each drug. For each drug, the DST results are indicated by up to 5 types of test conducted, namely bactec, hain, Le, GeneExpert and lpa (Galkina et al., 2012). In cases where the tests report different results for the same strain, we consider a cumulative DST case based on the profile reported by the majority of tests.

### Drug-interaction scores

For all combinations of drug regimens given to each patient, we computed a ‘drug-interaction score’ using the INDIGO-MTB tool (Ma et al., 2019), which uses Random forests to assign drug interaction scores that capture the nature of interaction between drugs using individual drug response transcriptomics data. The interactions are considered synergistic (score <0.9), additive (scores 0.9-1.2), and antagonistic (scores >1.2) respectively as used in prior studies

### Mtb families

Several Mtb families belonging to multiple lineages are represented in this dataset across different populations. The dataset reports Mtb strains from 27 different families in infected patients, with some patients infected with 2 or more families (which is unlikely). The TB portals database refers to these families as lineages, which is inaccurate. Lineage refers to the M. tuberculosis classification based on Large sequence polymorphism (LSP) or Single nucleotide polymorphisms (SNP). Family/sub-lineages refer to the classifications based on spoligotyping.

### Chest X rays and CT data

Chest X rays were available for a subset of the patients, taken across multiple time points, with Day 0 considered as the time TB infection was confirmed and treatment was initiated for the patient. Manual annotations of these X rays performed by radiologists and/or general physicians were made available. We chose the annotation information for X Rays recorded at day 0, and in instances where day 0 was not available, we chose the day closest to Day 0 to capture the early days of infection before treatment takes effect. Similarly, we chose the CT annotations closest to day 0 when available.

### Outcomes

The outcomes of infection for each patient was considered as a response variable in our analysis. Outcomes of infection and treatment are described for each patient, with 6 outcomes provided, namely *Cured, Completed, Failure, Died, Palliative Care, Lost to Follow up, Still on treatment and Unknow*n. As we are interested in analyzing successful and unsuccessful treatment outcomes, we pooled the outcomes ‘Cured’ and ‘Completed’ as ***“Success”***, while outcomes ‘Failure’, ‘Died’ and “Palliative Care” were grouped together under the outcome ***“Failure”***, as they all indicate unfavorable treatment outcomes per patient. We did not consider patients with outcomes *Lost to Follow up, Still on treatment and Unknow*n for our analysis.

### Data imputation

Initially, complete case analysis was performed with only patients that had less than 50% NaN values across all features. We repeated the analysis by imputing missing values using the K-nearest neighbor (KNN) imputation method, with k set to 3. This method was chosen as nearest neighbor imputation methods have been shown to be effective for machine learning with missing data for EHR analysis (Beaulieu-Jones and Moore, 2017).

### Data modalities

The data was split into 3 categories for analysis describing different aspects of host and pathogen data. These categories were a) patient social, clinical and demographic features b) patient radiological features as determined by chest X rays (CXR) and CT scans and c) pathogen genomic features which include gene mutations and drug sensitivity analyses.

### Machine Learning

We randomly split the input data into training (80%) and test (20%) samples for each category. The training data was then used to build a Random forest model to determine outcomes of Success and Failure. The function uses RandomizedGridSearch within cross-validation to tune the XGBoost hyperparameters and estimate model performance. The final model uses the hyperparameters from the best-scoring CV fold and is trained on the entire dataset. To account for the class imbalance in the two outcomes of success and failure, we considered an inverse weighting approach to balance the two classes (e.g. if success represents 80% of the outcomes, weights assigned are 1/0.08 and 1/0.02 for success and failure respectively). We performed a 5-fold cross validation and assessed model performance in predicting disease outcome on the training data. The model performance was further evaluated on the test dataset containing the remaining 20% data. Different metrics were considered to evaluate model performance including Accuracy, Precision, Recall, F1 score, Matthews Correlation coefficient and the Pearson’s R, to predict the outcome of infection. We conduct a random permutation analysis of the data to compute Feature selection was performed after evaluating for variables that account for >95% of the variance observed in the model based on their Cumulative scores for each feature. After identifying the top features for each category, we further evaluated the associations of different levels within the feature with the outcome by calculating p-values based on the hygecdf function (hypergeometric cumulative distribution)(Vidakovic, 2011) in MATLAB. Additional validation was performed on newer data populated in the TB portals database (January 2022).

### Statistical analysis

Pairwise statistical tests were performed for all features in the dataset. For comparing two continuous variables, the Kendall rank correlation was used, which is a non-parametric alternative to Pearson’s correlation(Akoglu, 2018). For two categorical variables, the Chi-squared test was performed to determine if there is a significant difference between the observed and expected frequencies of the associated contingency table. When comparing continuous and categorical variables, the Kruskal-Wallis test was used, which tests whether there is a difference between the groups of the continuous feature. The p-values from all of these tests were combined into a matrix and plotted as a heatmap. For all comparisons, a bonferroni correction test(Ranstam, 2016) was applied.

### Data Visualization

We use the Python Shapley package for data visualization. All machine learning and statistical analysis was conducted in Python version 3.7.13 and repeated in Matlab version R2021b.

## Data Availability

All data produced in the present work are contained in the manuscript

## Author contributions

AS and SC conceptualized the study, gained access to the NIAID TB portals data, analyzed the data and wrote the manuscript. AS, KS, CC, HSA conducted statistical and machine learning analyses for the data and prepared the figures. ZH, PA and SC supervised the study, designed the research and edited the manuscript. All authors read, contributed to, and approved of the manuscript.

## Code availability

The code for machine learning and statistical analysis conducted in this study is available on Synapse: https://www.synapse.org/#!Synapse:syn32889400

## Color legend

Blue - chest imaging features significantly associated with both resistance and treatment failure; Orange - Imaging features significantly associated only with failure; Gray - Imaging features associated significantly with drug resistance (p<0.05)

## Supplementary Information

**Supplementary Figure 1.**
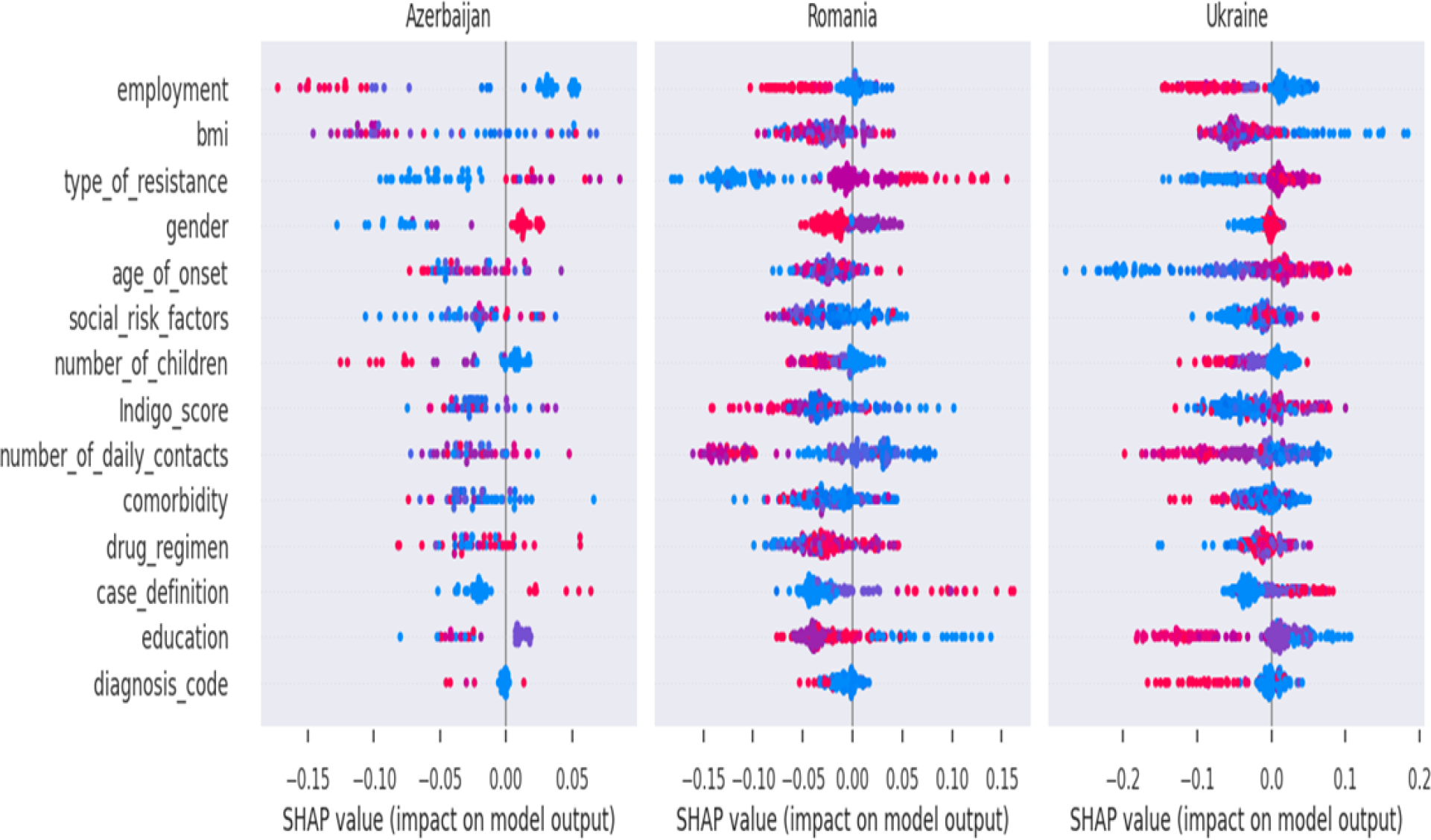
Shap feature value distributions and their impact on modeling Failure across countries Azerbaijan, Romania and Ukraine (the next set of populated countries after Moldova, Georgia and Belarus (Fig 3g). Each dot represents a single patient, and the colors range from blue (lower values) to red (higher values).

**Supplementary Figure 2.**
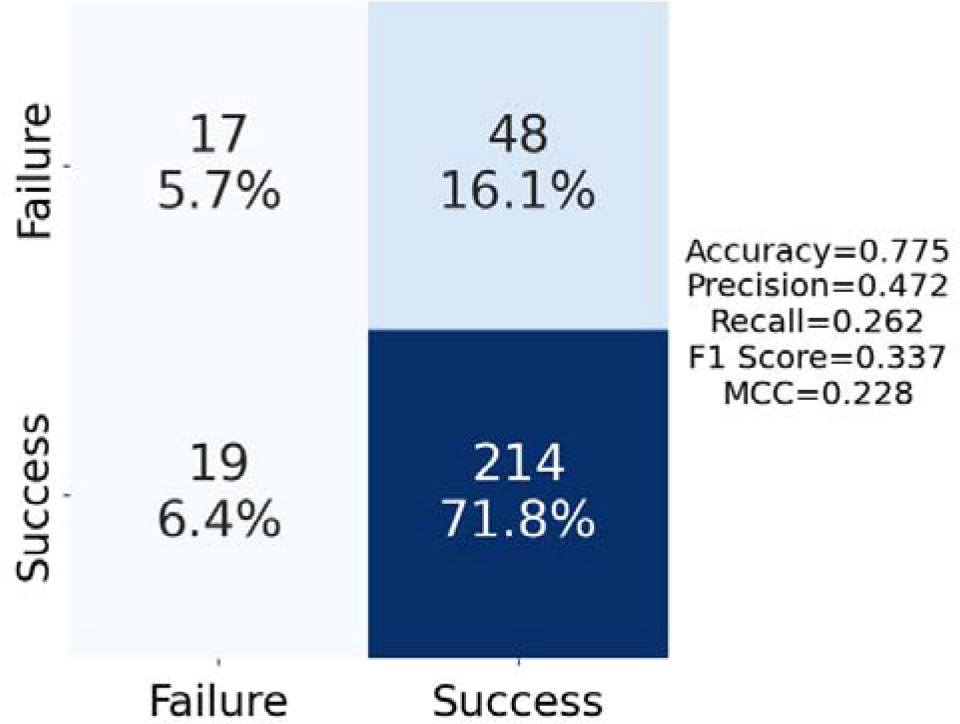
External validation of CXR modeling on new patient data

**Supplementary tables (Supplementary_tables.xlsx). Table S1**. MTB families seen in the data. **Table S2**. Drug-interaction FIC scores computed by INDIGO-MTB for all drug combinations in treatment regimens. **Table S3**. Feature associations among top features across all modalities with FDR corrected p-values.

